# The Use of Machine Learning Models and Radiomics for Segmentation and Classification of Adnexal Masses on Ultrasound: A multi-cohort retrospective study

**DOI:** 10.1101/2023.04.26.23289150

**Authors:** Jennifer F Barcroft, Kristofer Linton-Reid, Chiara Landolfo, Maya Al Memar, Nina Parker, Chris Kyriacou, Maria Munaretto, Martina Fantauzzi, Nina Cooper, Joseph Yazbek, Nishat Bharwani, Sa ra Lee, Ju Hee Kim, Dirk Timmerman, Joram M. Posma, Luca Savelli, Srdjan Saso, Eric O. Aboagye, Tom Bourne

## Abstract

**Background:** Ovarian cancer remains the deadliest of all gynaecological cancers. Ultrasound-based models exist to support the classification of adnexal masses but are dependent on human assessment of features on ultrasound. Therefore, we aimed to develop an end-to-end machine learning (ML) model capable of automating the classification of adnexal masses.

**Methods:** In this retrospective study, transvaginal ultrasound scan images were extracted and segmented from Imperial College Healthcare, UK (ICH development dataset; n=577 masses; 1444 images) and Morgagni-Pierantoni Hospital, Italy (MPH external dataset; n=184 masses; 476 images). Clinical data including age, CA-125 and diagnosis (ultrasound subjective assessment, SA) or histology) were collected. A segmentation and classification model was developed by comparing several models using convolutional neural network-based models and traditional radiomics features. Dice surface coefficient was used to measure segmentation performance and area under the ROC curve (AUC), F1-score and recall for classification performance.

**Findings:** The ICH and MPH datasets had a median age of 45 (IQR 35-60) and 48 (IQR 38-57) and consisted of 23·1% and 31·5% malignant cases, respectively. The best segmentation model achieved a dice surface coefficient of 0·85 ±0·01, 0·88 ±0·01 and 0·85 ±0·01 in the ICH training, ICH validation and MPH test sets. The best classification model achieved a recall of 1·00 and F1-score of 0·88 (AUC 0·93), 0·94 (AUC 0·89) and 0·83 (AUC 0·90) in the ICH training, ICH validation and MPH test sets, respectively.

**Interpretation:** The ML model provides an end-to-end method of adnexal mass segmentation and classification, with a comparable predictive performance (AUC 0·90) to the published performance of expert subjective assessment (SA, gold standard), and current risk models. Further prospective evaluation of the classification performance of the ML model against existing methods is required.

**Funding:** Medical Research Council, Imperial STRATiGRAD PhD programme and Imperial Health Charity.

**Research in Context:** *Evidence before this study:* Adnexal masses are common, affecting up to 18% of postmenopausal women. Ultrasound is the primary imaging modality for the assessment of adnexal masses. Accurate classification of adnexal masses is fundamental to inform appropriate management. However, all existing classification methods are subjective and rely upon ultrasound expertise. Various models have been developed using ultrasound features and serological markers such as the Risk of malignancy index (RMI), International Ovarian Tumour Analysis (IOTA) Simple Rules (SR), the IOTA Assessment of Different NEoplasia’s in the AdneXa (ADNEX) model, and American College of Radiology (ACR) Ovarian-Adnexal Reporting and Data System Ultrasound (ORADS-US) to support the classification of adnexal masses. Despite modelling efforts, expert subjective assessment remains the gold standard method of classifying adnexal masses. The use of machine learning (ML) within clinical imaging is a rapidly evolving field due to its potential to overcome the subjectivity within image assessment and interpretation. Various studies (n=17) evaluating the use of ML within the classification of adnexal masses on ultrasound have been summarised within a recent meta-analysis by Xu et al, 2022. No studies used a radiomics-based approach to the classification of adnexal masses, and most have not been externally validated within a test set, questioning their generalisability. The largest study to date (Gao et al, 2022), used a deep learning (DL) based approach and was externally validated, yet its performance (F1 score 0·551) was not comparable to existing classification approaches.

*Added value of this study:* We have developed an end-to-end ML model (ODS) using DL and radiomics-based approaches, capable of identification (automated segmentation) and classification of adnexal masses with a high detection rate for malignancy. The ODS model had a performance comparable to the published performance of existing adnexal mass classification methods and does not rely upon ultrasound experience.

*Implications of all the available evidence:* ODS is a high performing, end-to-end model capable of classifying adnexal masses and requires limited ultrasound operator experience. The ODS model is potentially generalisable, having showed consistent performance in both validation (internal) and test (external) sets, highlighting the potential clinical value of a radiomics-based model within the classification of adnexal masses on ultrasound. The ODS model could function as a scalable triage tool, to identify high risk adnexal masses requiring further ultrasound assessment by an expert.

## Introduction

Ovarian Cancer (OC) affects 2% of women in their lifetime and remains the leading cause of death from a gynaecological malignancy in the UK(1). The poor prognosis of OC is mainly attributed to most women (75%) presenting late, with advanced stage disease(1). Unfortunately, an effective OC screening program does not exist; therefore, diagnosis is reliant upon prompt recognition of gynaecological symptoms and accurate interpretation of clinical imaging(2).

Adnexal masses are common, affecting up to 18% of postmenopausal women in the UK (3). The accurate classification of adnexal masses is fundamental to ensure malignant adnexal masses are promptly identified and undergo surgical intervention by an appropriately trained surgeon. Particularly in younger women and those with an asymptomatic lesion, it is important that those with a benign mass are not subjected to unnecessary intervention, with potential complications. (4).

Expert subjective assessment (SA) is the gold standard method for classifying adnexal masses, yet is restricted by the availability of expert examiners(5). Various ultrasound-based diagnostic models, using a combination of ultrasound features with or without serological markers (CA-125) exist to support the classification of adnexal masses, including the Risk of malignancy index (RMI)(6), International Ovarian Tumour Analysis (IOTA) Simple Rules (SR) (7), the IOTA Assessment of Different Neoplasia’s in the AdneXa (ADNEX) model (8) and American College of Radiology (ACR) Ovarian-Adnexal Reporting and Data System Ultrasound (ORADS-US)(9).

ADNEX is the best performing ultrasound-based model with an AUC of 0·94, sensitivity of 85·2% (fixed specificity 90%) and specificity of 85·7% (fixed sensitivity 90%). RMI is the recommended approach within UK National Guidance for the assessment of adnexal masses in post-menopausal women(6). RMI has an AUC of 0·89, sensitivity of 70·1% (fixed specificity 90%) and specificity of 69·3% (fixed sensitivity 90%) (10). However, expert SA remains the best method for classifying adnexal masses, with an AUC of 0·96, sensitivity of 0·90 and specificity of 0·91(5). Although ultrasound-based models have been internally and externally validated in the hands of expert and non-expert ultrasound examiners, their use involves the subjective interpretation of ultrasound-based features (5,11,12).

Machine learning (ML) refers to the enablement of computers to perform tasks without the need for explicit programming. Deep Learning (DL) is a subset of ML, that specifically refers to the use of artificial neural networks, which has shown great promise within clinical image analysis(13). Typically, DL models consist of a series of interconnecting neural networks and nodes, which form connections through a process of supervised training, involving repeated exposure to adnexal mass images, linked to the clinical/histological diagnosis(13). Radiomics broadly defines a process which involves extracting high-throughput quantitative features from images. In particular, first order statistics including the median and mean pixel/voxel intensity and higher order features such as textural features, wavelet and functions of wavelet transformed images(14).

Given the differences in the morphology between benign and malignant adnexal masses on ultrasound, we would anticipate a similar discrete pattern in the radiomics features (signature) which could be utilised to develop a ML classification model. A radiomics-based model provides a degree of mathematical explainability to the classification output, which is invaluable within clinical image interpretation (15).

Current methods of adnexal mass classification rely on ultrasound experience and are prone to error. There is a need for a robust ML model, capable of classifying adnexal masses, which does not rely upon prior ultrasound experience, and can provide a scalable, generalisable, accurate solution to adnexal mass classification.

There has been recent interest in the integration of ML into the ultrasound classification of adnexal masses(16). A recent meta-analysis and systematic review highlighted 17 studies applying ML models (principally DL) in this field(16). The pooled model performance had a sensitivity of 0·91 and specificity of 0·87 (16). The largest ML study by *Gao et al* outlined the development and external validation of a DL-based model to automate the classification of adnexal masses. In the validation dataset, the model had an AUC of 0·911 and F1-score of 0·812, whereas in an external test set, the model had an AUC of 0·870, but an F1-score of 0·551(17). The F1-score adjusts for class imbalance (true negatives do not contribute to the score) and so is the performance metric of choice when evaluating an ML model, rather than AUC.

Our study aims to extend our previous work in Computed Tomography (CT) scanning, which focused on the development and validation of a radiomics-based model to improve the prediction of the prognosis for women with OC (18,19). We aim to develop and externally validate a robust ML model, utilising both radiomics and DL approaches capable of classifying adnexal masses on ultrasound. In addition, we shall determine the value of integrating various clinical parameters such as CA-125 and age on the classification performance of the model. Finally, recognising the importance of a robust, generalisable model its performance will be evaluated in an external test set of adnexal masses.

## Methods

### Study design and participating cohorts

This retrospective study consisted of women (≥18 years) recruited from two European Gynaecology Oncology centres, between December 2017 and September 2022: (1) Imperial College Healthcare NHS Trust, London, UK (ICH) and (2) Morgagni-Pierantoni Hospital, Forli, Italy (MPH). The ML model was developed and internally validated on adnexal mass images from ICH (Development n=577 masses; 1444 images) and externally validated utilising the MPH dataset (test n=184 masses; 476 images).

Eligibility criteria included: a non-physiological adnexal mass which had been: (1) expectantly managed for six months and classified as benign according to ultrasound SA or (2) undergone surgical removal with available histology. Exclusion criteria included: no transvaginal ultrasound images, physiological cysts, and cases with only ‘split screen’ images. Pregnancy was not an exclusion criterion.

### Ultrasound image acquisition

All ultrasound examinations were carried out using a standardised approach, with the application of IOTA terms and definitions(20). Subjective assessment (SA) was used to classify the adnexal mass(es) as benign or malignant. Adnexal masses were scanned on various ultrasound systems including: Voluson GE (Voluson E6, E8, E9, E10, P8, S10), Samsung (W10, WS80, HS60) and Esaote (mylab). 2D ultrasound, grayscale adnexal mass images without any callipers were taken for each adnexal mass. The ultrasound examinations were carried out by ultrasound examiners of varying experience, but under expert supervision. The pseudo-anonymised grayscale adnexal mass images were exported as TIFF images. Adnexal masses which required surgery because of symptoms, suspicion of malignancy or patient choice were removed by an appropriately trained surgeon within the centres. The World Health Organisation (WHO) classification was used to define the tumour sub-type and the International Federation of Gynaecology and Obstetrics criteria was used to stage malignant adnexal masses (21). Clinical data including age and CA-125 level was also collected where available.

### Ethical approval

This study was approved by the UK Regional Ethics Committee (05/QO406/178). All procedures involving human participants were in accordance with the ethical standards of the institutional and/or national research committee and with the principles of the 1964 Declaration of Helsinki and its later amendments or comparable ethical standards. The use of anonymous external dataset images was granted by Imperial College Ethics Committee (22IC7780).

### Model development

The region of interest was defined in a process known as segmentation within 3D Slicer (https://www.slicer.org/), an open-source segmentation software by an experienced ultrasound operator (J·F·B) (22). The segmented images were checked individually by a level III (expert) ultrasound examiner (C·L, M·M, S·S, N·B)(23). In accordance with the Image Biomarkers Standardization Initiative (IBSI) scans were resampled to isotropic 1×1mm2(24). All continuous variables were scaled and mean centred using training data set statistics.

To enable the development of an end-to-end classification model, the first step involves the development of a segmentation model, to identify the region of interest (lesion), the second step requires the development of a classification model to determine if the lesion is benign or malignant.

We developed several convolutional neural network (CNN)-based segmentation models, through training on the ICH development training dataset (462 masses, 1155 images). The segmentation performance was evaluated on (1) ICH validation (115 masses 289 images) (2) MPH test set (184 masses, 476 images) (Table 3).

**Table 1:** Summary of the characteristics of the ICH Development (training and internal validation) and MPH External test set: patient demographics, presence of histological diagnosis and CA-125 level. The differences between individual parameters are demonstrated with the respective p values.

| Parameter |  | ICH Development<br>n= 577, 1444 images | MPH Test<br>n=184, 476 images | P-value |
| --- | --- | --- | --- | --- |
| Age: median (IQR) years |  | 45 (35-60) | 48 (38-57) | 0.63 |
| Number of images<br>(median, IQR) |  | 2 (2-3) | 2 (2-3) |  |
| Diagnosis | Benign | 444 (76.9%) | 126 (68.5%) | 0.029 |
|  | Malignant | 133 (23.1%) | 58 (31.5%) |  |
| Histology (%) | Yes | 285 (49.4%) | 184 (100.0%) | $2.22 \times 10^{-16}$ |
|  | No | 292 (50.6%) |  |  |
| CA-125 U/ML (median, IQR) |  | 25 (14-114) | 16.5 (10-54.8) | 0.062 |

**Table 2:** Adnexal mass diagnosis, based on histology or ultrasound (expert subjective assessment) within the ICH development (training and internal validation) and MPH external test set (n, %). There was a significant difference in adnexal mass sub-types between the two datasets (ICH, MPH), demonstrated with a p value of 0·000000027.

| **Type of adnexal mass** | **N, %**<br>**ICH Development**<br>**(training and**<br>**validation)** | **N % MPH Test Data** |
| --- | --- | --- |
| Cystadenoma (serous, mucinous, seromucinous) | 179 (31.0%) | 44 (23.9%) |
| Dermoid | 100 (17.3%) | 31 (16.8%) |
| Endometrioma | 56 (9.7%) | 33 (17.9%) |
| High Grade Serous Carcinoma | 41 (7.1%) | 8 (4.3%) |
| Cystadenofibroma | 40 (6.9%) | 4 (2.2%) |
| Benign tubal | 31 (5.4%) | 0 (0.0%) |
| Serous Borderline | 21 (3.6%) | 15 (8.2%) |
| Mucinous Borderline | 17 (2.9%) | 1 (0.5%) |
| Fibroma | 18 (3.1%) | 8 (4.3%) |
| Benign Other | 15 (2.6%) | 5 (2.7%) |
| Endometroid ovarian cancer | 10 (1.7%) | 2 (1.1%) |
| Clear cell ovarian cancer | 7 (1.2%) | 2 (1.1%) |
| Sex cord | 7 (1.2%) | 10 (5.4%) |
| Struma Ovarii | 5 (0.9%) | 0 (0.0%) |
| Metastasis | 5 (0.9%) | 4 (2.2%) |
| Mucinous Carcinoma | 4 (0.7%) | 6 (3.3%) |
| Malignant other | 4 (0.7%) | 2 (1.1%) |
| Carcinosarcoma | 6 (1.0%) | 3 (1.6%) |
| Seromucinous Borderline | 6 (1.0%) | 6 (3.3%) |
| Germ cell | 3 (0.5%) | 0 (0.0%) |
| Other Borderline (Brenner, Endometroid) | 2 (0.3%) | 0 (0.0%) |
| TOTAL | 577 (100.0%) | 184 (100.0%) |

**Table 3:** Performance of Ovarian Diagnostics score (ODS) radiomics model, DL models and CA-125 baseline model in the ICH Training, ICH validation and MPH external test set. Performance metrics summarised: F1 score (F1), Area under the ROC curve (AUC), Precision (PREC), Recall (REC), and Specificity (SPEC).

| Model | ICH TRAINING |  |  |  |  | ICH VALIDATION |  |  |  |  | MPH TEST |  |  |  |  |
| --- | --- | --- | --- | --- | --- | --- | --- | --- | --- | --- | --- | --- | --- | --- | --- |
| Metrics | F1 | AUC | PREC | REC | SPEC | F1 | AUC | PREC | REC | SPEC | F1 | AUC | PREC | REC | SPEC |
| ODS | 0.88 | 0.93 | 0.78 | 1.00 | 0.80 | 0.94 | 0.89 | 0.89 | 1.00 | 0.80 | 0.83 | 0.90 | 0.72 | 1.00 | 0.73 |
| ResNet-18 | 0.93 | 0.97 | 0.93 | 0.93 | 0.95 | 0.81 | 0.85 | 0.85 | 0.77 | 0.79 | 0.75 | 0.80 | 0.82 | 0.78 | 0.83 |
| ResNet-34 | 0.92 | 0.97 | 0.91 | 0.93 | 0.93 | 0.76 | 0.81 | 0.82 | 0.80 | 0.77 | 0.77 | 0.80 | 0.83 | 0.77 | 0.84 |
| ResNet-50 | 0.88 | 0.96 | 0.91 | 0.84 | 0.93 | 0.37 | 0.61 | 0.41 | 0.34 | 0.35 | 0.33 | 0.54 | 0.40 | 0.29 | 0.28 |
| CA-125 | 0.11 | 0.49 | 0.08 | 0.18 | 0.10 | 0.36 | 0.67 | 1.00 | 0.22 | 0.99 | 0.38 | 0.49 | 0.33 | 0.45 | 0.35 |

Radiomics features were extracted using TexLab 2·0, with various grey-level binning ranges, to compute a total of 3906 radiomics based features. We explored a combination of 8 different feature reduction techniques with 11 different ML algorithms, including linear and tree-based techniques, and boosted and regularised variations. Feature reduction methods included mutual information, recursive feature elimination, correlation-based and linear methods. Hyper-parameter optimisation was performed via grid-search with ten-fold cross-validation optimising for concordance index. Hyper-parameter ranges of the models are listed in Supplementary Table 1. The binarization of predicted probabilities (malignant vs benign) was derived from a threshold of 0·5. A description of algorithms and feature reduction methods are detailed in the Supplementary Material.

In addition to traditional radiomics features, we also built several CNN classification models. ResNet-based DL architectures were applied without the segmentation region of interest. ResNet-18,-32, and -50.

We used the Radiomics Quality Score (RQS) (25) and Transparent Reporting of a Multi-variable Prediction Model for Individual Prognosis or Diagnosis (TRIPOD, https://www.tripod-statement.org/) guidelines for reporting the development and validation of the prediction models.

### Outcomes

The primary outcome of the segmentation ML model was identification of the region of interest (ROI), compared to ultrasound examiners segmentation (ground truth). The primary outcome of the classification ML model was adnexal mass diagnosis (benign or malignant), compared to ultrasound SA or histological diagnosis.

### Data availability

The trained automated segmentation models and code are available at wwwe.github.com/XXX. The anonymised adnexal image datasets used in this study are not available as open-source datasets.

### Statistical analysis

To assess the model’s performance for the accurate classification of an adnexal mass, we used the F1-Score (harmonic mean of precision and recall), precision and recall. For clinical relevance we calculated AUC, sensitivity, and specificity. We assessed the calibration of the ODS model, through evaluation of the calibration intercept and slope for ICH training, ICH validation and MPH test set. The purpose of calibration was to determine whether the ODS model over or underpredicted the risk. Quantitative statistics were presented as median and interquartile range (IQR). Continuous variables were compared using Wilcoxon signed-rank tests, and categorical variables were compared using the Fisher’s exact test. Statistical analysis of clinical variables was two-sided, and Benjamini–Hochberg multiple testing corrected p values of less than 0·05 were used to indicate statistical significance. Calibration curve statistics were computed using the Regression Modelling Strategies package version 6·5, using the non-parametric confidence intervals method described by Qin and Hotilovac(26). With the single variable CA-125 model, we performed a univariate logistic regression. Missing CA-125 values were imputed using the multivariate imputation by chained equations algorithm.

## Results

The ICH development dataset consisted of 577 cases (1444 images); the median age was 45 (IQR 35-60). All malignant cases (23·1%) were managed surgically and high-grade serous carcinoma (n=41, 7·1%) was the commonest malignant adnexal mass. Most benign cases were managed conservatively (n=292, 65·8%) and cystadenoma (n=179, 31·0%) was the commonest benign mass. Serum CA-125 levels were available for 301 cases (52·2%), median value of 25 U/ml (IQR 14-114).

The MPH test dataset consisted of 184 cases (476 images); the median age was 48 (IQR 38-57). All MPH cases were managed surgically, with a malignancy rate of 31·5%, which was significantly higher than ICH (p=0·029). CA-125 data was available for 108 cases (58·7%), median value 16·5 U/ml (IQR 10-54·8). Serous borderline (n= 15, 8·2%) and cystadenoma (n=44, 23·9%) were the most common malignant and benign adnexal masses respectively. The difference in adnexal sub-types between the ICH and MPH dataset were statistically significant (p<0·001). The MPH test data set contained 150 images (31·5%) with callipers present, we found that the presence of callipers did not contribute to the explained variance within the data set (supplementary Figure 4).

The best performing segmentation model, utilised DL, achieved a dice surface coefficient (SD) of 0·85 ± 0·01, 0·88 ±0·01 and 0·85 ± 0·01, for ICH training, ICH validation, and MPH test set respectively (Figure 3). The best performing classification model, at a threshold of 0·5, termed the Ovarian Diagnostic Score (ODS), utilised ridge regressions with Pearson correlation-based feature reduction (Figure 4). The ODS model reached an F1-score of 0·88 (AUC 0·93) in the ICH training, 0·94 (AUC 0·89) in ICH validation and 0·83 (AUC 0·90) in the MPH external test set.

**Figure 1.**
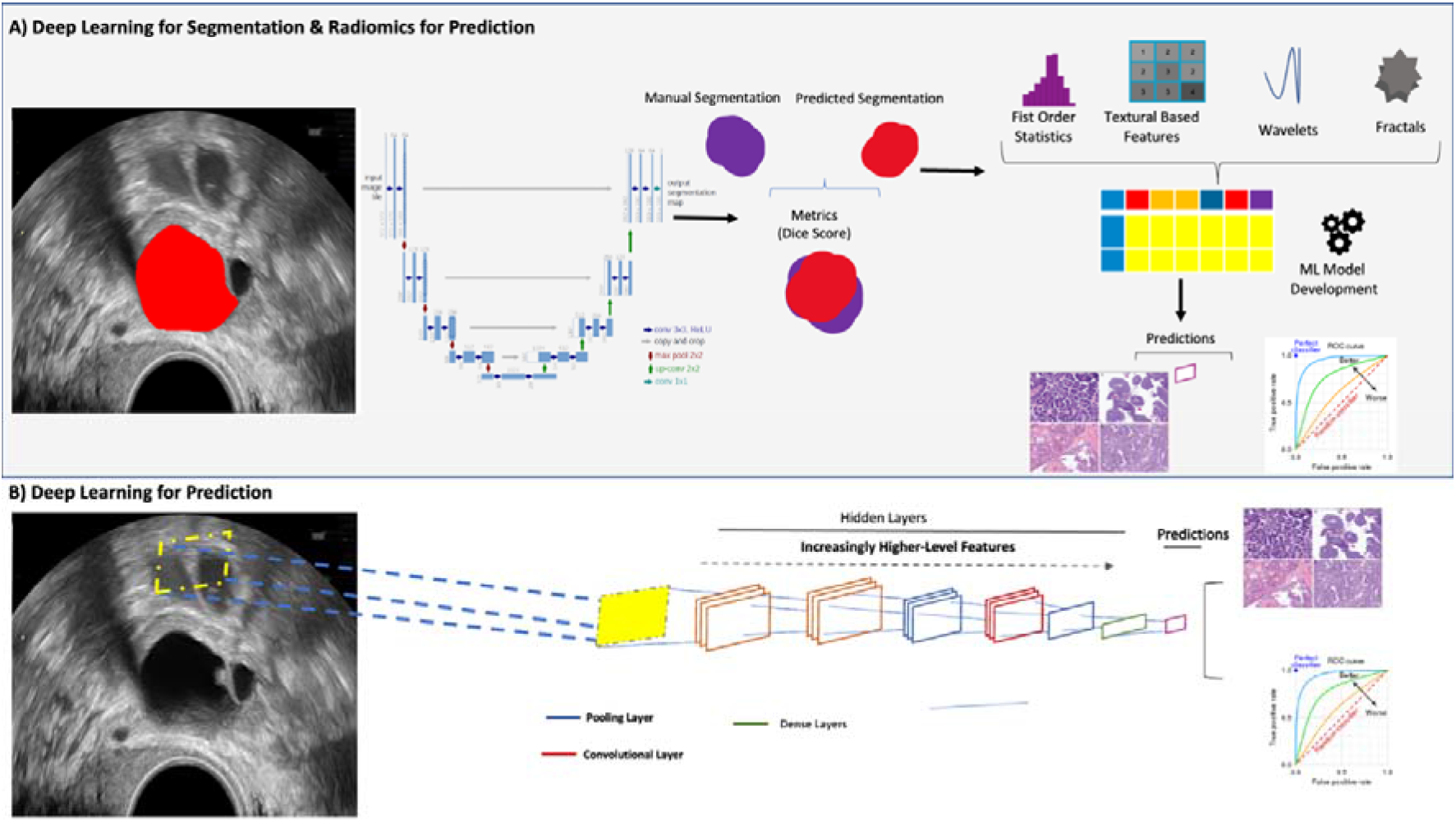
Overview of AI Approach. An end-to-end approach can either rely on a combination of Deep Learning (DL) for segmentation and radiomics, or a direct DL-based approach without the need for segmentation. **(A)** The traditional radiomics feature pipeline from segmented region of interest to radiomics feature computation and ML modelling. **(B)** DL approach, using convolutional neural networks, to facilitate auto-segmentation and classification.

**Figure 2:**
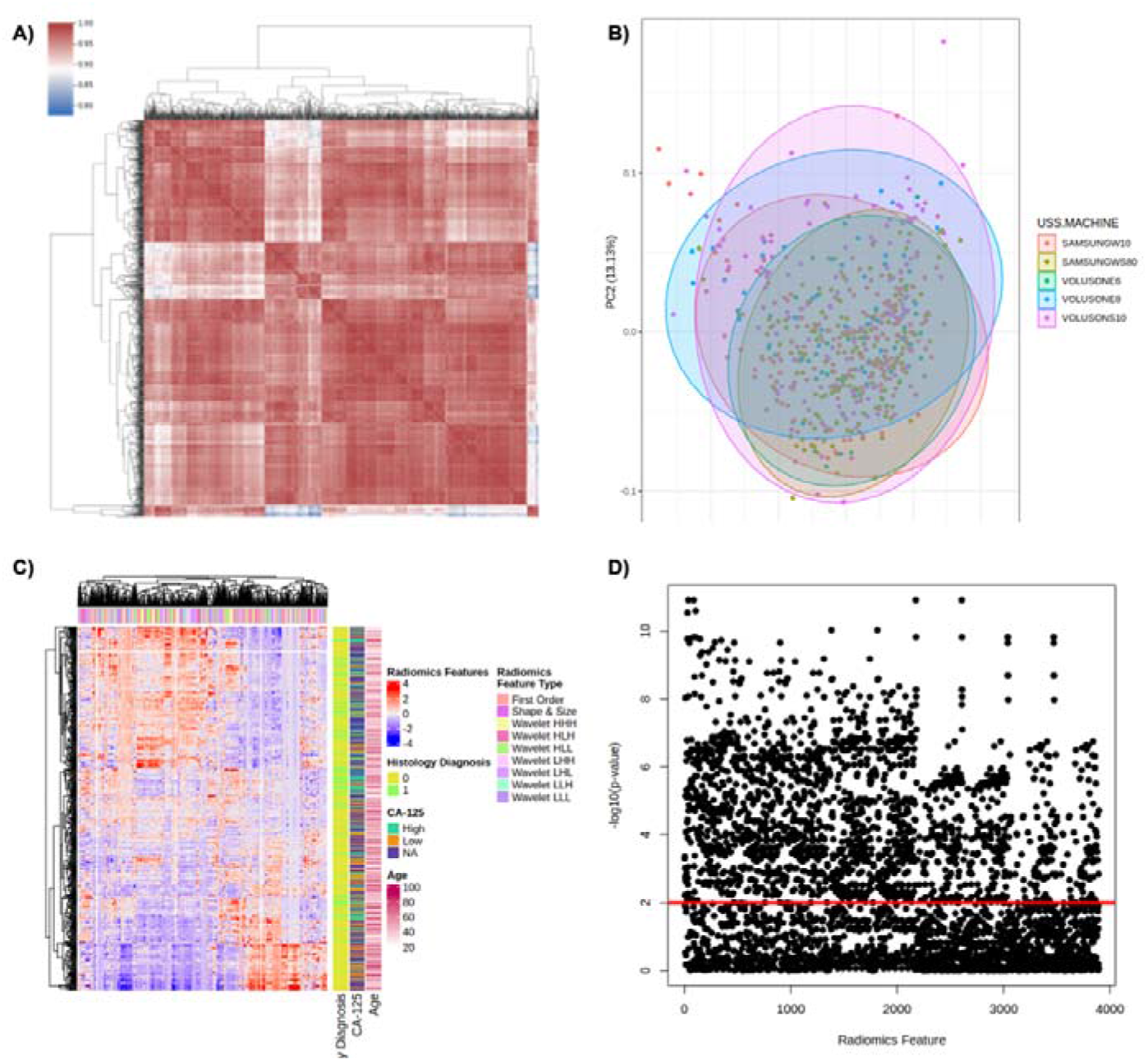
Overview of Radiomics data structure. (A) Correlation matrix heat map of radiomics parameters. The degree of correlation between radiomics parameters is indicated within the heatmap (red indicates perfect correlation). (B) Principal component analysis (PCA) indicates the degree of explained variance within the dataset (ultrasound scanner type), PC1 and PC2 explain 37·6% and 13·14% of explained variance. (C): Heatmap of all extracted radiomics features for adnexal mass classification. Each row corresponds to an individual patient and each column corresponds to each scaled radiomics feature. The colour key outlines the corresponding radiomics feature sub-type. Clinical parameters including age and CA-125 are represented. (D) Univariate logistic regression outlining radiomics features and their respective univariate logistic regression derived p-values (horizontal red line indicates p<0·01)

**Figure 3:**
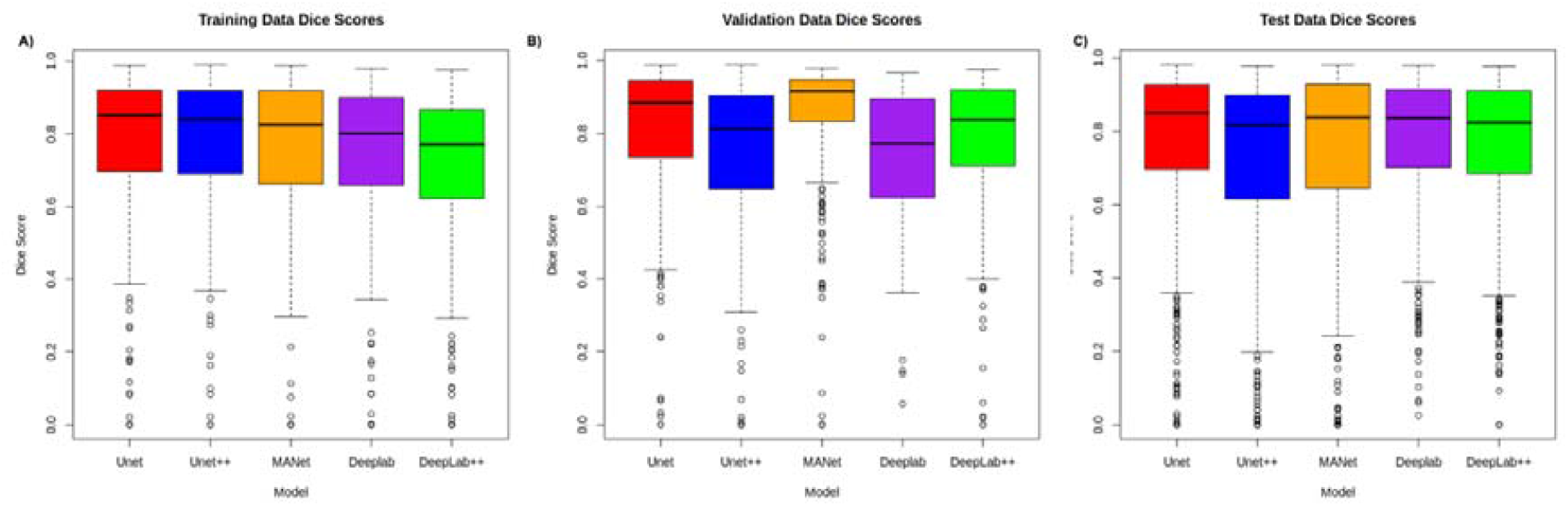
The performance of five DL segmentation models: (1) Unet (2) Unet++ (3) MANet (4) Deeplab (5) Deeplab++. compared to ground truth segmentation within the A: Training dataset, B: Validation dataset C: Test dataset. The similarity scores (dice surface coefficient, Dice scores), presented within a box plot, the middle line corresponds to the median, the upper and lower boundaries of the box correspond to upper and lower quartiles, whilst the whiskers reflect the minimum and maximum value and the white dots below the whiskers correspond to outliers.

**Figure 4:**
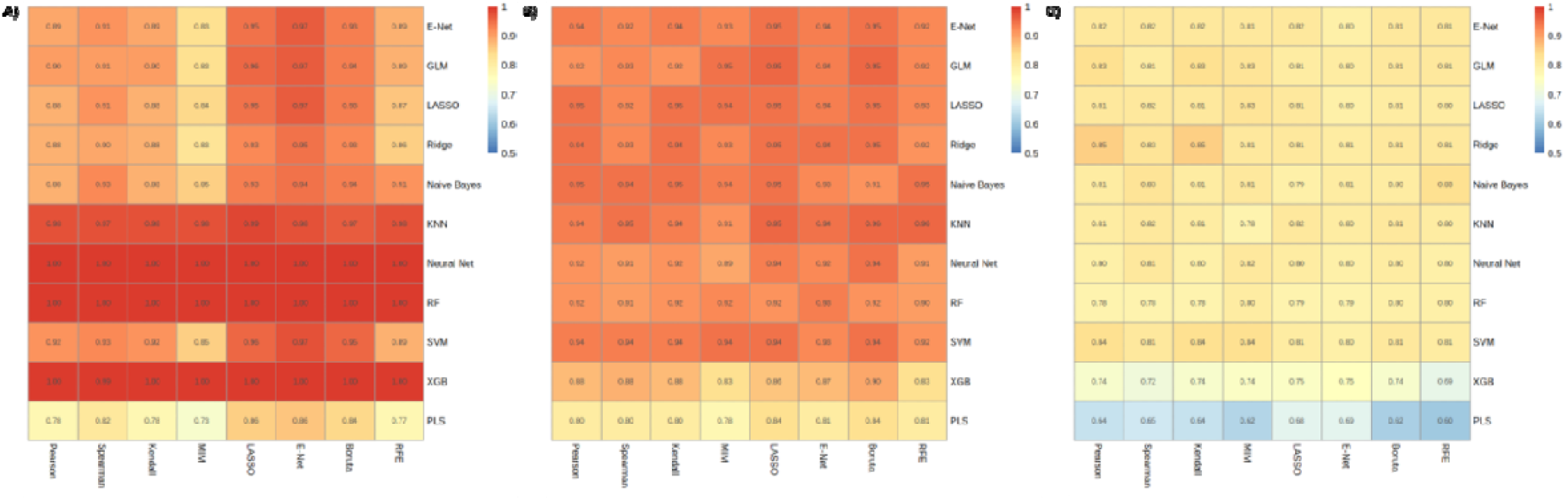
Supervised Radiomics Modelling. In heatmaps, x-axis corresponds to feature selection techniques and y-axis to modelling strategy; values are F1-scores. Plots (A)-(C), represent ICH training, ICH validation, and MPH test set results, respectively.

**Figure 5:**
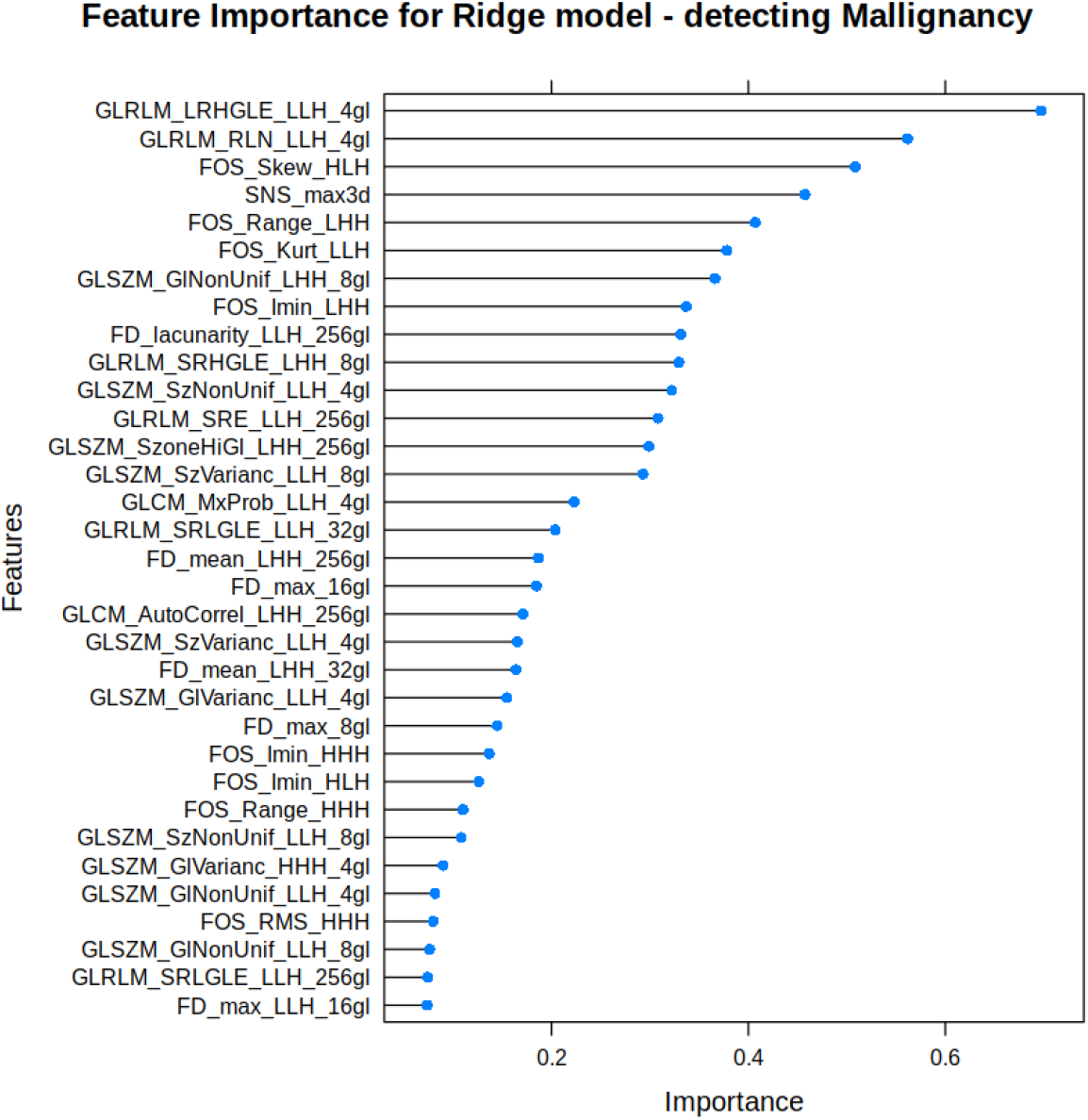
Feature Importance of Ridge Regression Model for the prediction of malignancy.

The ODS model, outperformed the CA-125-based model, in the ICH validation (F1 score 0·94 vs. 0·36) and MPH test (F1 score 0·83 vs. 0·38) respectively. The ODS model had a recall of 1·00 in the ICH training, ICH validation and MPH test sets respectively, so there were no false negatives cases. The ODS model had a precision (PREC) of 0·78 in ICH training, 0·89 in ICH validation and 0·72 in the MPH test set, translating to a false positive rate of 28% in MPH, compared to 11% in ICH validation. The specificity (SPEC) of ODS model was 0·80 in both the ICH training and validation, and 0·73 in the MPH test set.

The calibration curve for the ODS model had an intercept of 0·00000037, -0·87, -0·55 in the ICH training, ICH validation, MPH test set respectively (a perfect intercept is 0). The slope of the calibration curve evaluates the spread of the estimated risks (target value of 1). The model had a slope of 1·39, 1·06, 0·25 in the ICH training, ICH validation and test set respectively. The slope of the MPH test indicates that the estimated risks are too extreme, i.e., higher predicted probability for high-risk patients and lower predicted probability for low-risk patients, compared with the ICH validation dataset (with more moderate risk estimates). The over-prediction (false positive) pattern seen in the MPH test set, is reflected in the lower precision of 0·72, compared to 0·89 in the ICH validation (Supplementary Figure 1).

## Discussion

We have demonstrated that the ODS model provides an end-to-end, method of adnexal mass segmentation and classification, with comparable predictive performance (F1 0·83, AUC 0·90) to the published performance of expert SA (AUC 0·96) and the ADNEX model (AUC 0·94). This ODS model has potential clinically utility, through its ability to automate the identification of the region of interest and provide a real-time classification of an adnexal mass, without the need for prior ultrasound operator experience.

The detection of malignancy is fundamental to any diagnostic test. Prioritising the detection of positive cases (recall) is often at the expense of the specificity and can result in a high false positive rate (FPR). The ODS model had a recall (sensitivity) of 1·00 in the ICH validation and MPH test sets, translating to a high malignancy detection rate. The precision, also known as positive predictive value of the ODS model was 0·89 and 0·72 in the ICH validation and MPH test set respectively, corresponding to a FPR of 11 and 28%. Given the high sensitivity of the ODS model, it could have value as an initial triage tool (first step), to identify ‘high risk’ adnexal masses, which warrant further evaluation by an expert ultrasound examiner (second step). This two-step approach could reduce the workload on an expert examiner, through focusing on identifying ‘high risk cases’ and would also address the relatively high FPR related to utilising a ODS model independently. Further prospective validation of the ODS model is required given our relatively small dataset, to determine its performance against existing methods of adnexal mass classification, before it could be considered as a clinically useful diagnostic test.

This is the largest radiomics study of ultrasound classification of adnexal masses to date, incorporating 577 cases (1444 images) in the development dataset (ICH) and externally validated (MPH) on 184 test cases (476 images). To facilitate the application of the ODS model to all adnexal masses (including those selected for conservative management), the training set included expectantly and surgically (histology) managed adnexal masses (Table 1). A two-step segmentation pipeline was implemented, involving an expert review of each adnexal mass image to ensure the dataset is of the highest quality, whilst minimising operator bias.

The methodical model development pipeline involved the evaluation of eight feature reduction and 11 ML techniques in various combinations, to establish the best performing ML model (ODS model) for this classification task. The ODS model at a threshold of 0.5 performed well in both the ICH (validation, F1 0·94, AUC 0·89) and MPH (external, F1 0·83, AUC 0·90) test sets, demonstrating its generalisability and applicability. In addition to the radiomics models, we also developed ResNet based DL models for the classification of malignancies without the need for manual segmentation. Whilst ResNet based architectures performed well in training, there was a decrease in performance in the validation and test sets. This is a key indicator that these models were overfitting especially with the largest ResNet-50 architecture despite our augmentation efforts. Overfitting is a common occurrence in DL models; however, it is expected that with an increase in data set sizes this issue would be resolved.

Radiomics-based models offer a degree of explainability to the classification output. The radiomics features Grey Level Run Length Matrix (GLRLM) and a Grey Level Size Zone (GLSZM) dominated the feature space that defined ODS. GLRLM quantifies grey level runs that are the length in number of pixels, of consecutive pixels that have the same grey level value while GLSZM quantifies grey level zones that are the number of connected pixels that share the same grey level intensity. Both reflect heterogeneity and biophysically begins to define the distinguishing features of benign versus malignant adnexal masses (18).

Using a radiomics-based approach coupled with a diverse training set may explain the superior predictive performance of the ODS model within the ICH validation (F1 score: 0·94) and MPH test set (F1-score: 0·83), compared to the largest ML study to date by Gao et al, which used DL (F1-score: 0·551)(17), suggesting that a radiomics-based approaches may be more suited to smaller clinical datasets(17). The drop in the performance of the ODS model between our internal and external test sets, is expected given the heterogeneity of adnexal mass sub-types (p<0·01), malignancy rates (p<0·05), scanner types and geographically diverse locations between the validation and test datasets. Furthermore previous studies (27–29) have used AUC as the performance metric, rather than F1-score, which may have overestimated the respective model’s performance at the classification of adnexal masses, given the lack of adjustment for class imbalance. Several ML models published in the literature have not been evaluated in an external test set; thus, limiting the conclusions that can be drawn on their performance and overall generalisability (27,29,30,30).

Whilst overfitting and adnexal mass heterogeneity is an ongoing challenge and has been demonstrated with an observed drop in classification performance and calibration, our results demonstrate the potential benefit of an end-to-end model capable of triaging adnexal masses. Establishing a large multi-centre cohort will look to overcome the challenges associated with relatively small datasets and aim to improve the overall calibration of radiomics (ML) and DL models and their potential clinical translation.

The retrospective extraction of adnexal mass images for the development of the ML model limited the prospective application of ADNEX and RMI models to enable direct comparison and so we must be cautious when comparing possible performance. The performance of existing clinical models within the literature (RMI, SR and ADNEX) is evaluated using AUC. The ML model performance on the external test set, based on AUC of 0·90 (F1score 0·83), is comparable to RMI (AUC 0·89), but slightly inferior to expert SA (AUC 0·96) and ADNEX (AUC 0·94), respectively(10,11). Whilst the performance of the ML model has not surpassed the published performance of existing classification models, the ODS model does offer a potential end-to-end diagnostic approach, which does not require expertise in adnexal mass classification to be used.

Finally, prospective evaluation of the ML model’s ability to correctly classify adnexal masses against existing methods (SA, RMI, ADNEX and ORADS-US classification system) is required before we can establish its value as a potential diagnostic tool. Furthermore, evaluation across multiple centres, in the hands of operators of varying experience is important, to establish the generalisability and clinical utility of the ML model.

## Conclusion

We have developed a high performing automated end-to-end ML model, capable of accurately classifying adnexal masses, with a good malignancy detection rate. Subject to further external validation, the ML model may be widely applicable, given the consistent performance of the ODS model across both internal and external datasets. A ML model could offer a scalable, accurate triage tool to effectively identify cases deemed ‘high risk’ of malignancy, which would warrant further expert ultrasound evaluation. A model, which is not reliant upon ultrasound expertise to classify adnexal masses, could address inherent barriers limiting the use of existing ultrasound-based models. Further work is required to evaluate the performance of the ODS model prospectively against existing methods of adnexal mass classification (expert SA, RMI, SR, ORAD-US and ADNEX) to further establish its role within the adnexal mass classification pipeline.

## Data Availability

data is available upon reasonable request to the authors

## Author roles

J·F·B and K·L-R contributed equally to this study. J·F·B, K·L-R, T·B, E·O·A and S·S were responsible for the original study design, drafting and protocol revision. J·F·B, C·K, N·P and N·C were responsible for patient recruitment and ultrasound image extraction at ICH. J·H·K, S·L, L·S, M·M, M·F were responsible for the clinical data and ultrasound image extraction for the external test set. J·F·B was responsible for clinical data extraction and segmentation of the ultrasound images. C·L, M·A-M, N·B and S·S were responsible for expert review of segmented ultrasound images. K·L-R was responsible for the development and validation of ML models and statistical analysis aspect of the manuscript. E·Y-T, V·P·L, I·J·C and V·L were responsible for data extraction and statistical analysis aspect of the manuscript. J·F·B, K·L-R, T·B, J·P, E·O·A and S·S were involved in writing and preparation of the manuscript, including tables and figures. K·L-R, E·O·A and J·P (focus: Machine learning and statistical analysis) provided technical input into the development of machine learning models and addressed statistical-related issues within the manuscript. J·F·B, T·B, C·L, M·A-M, J·Y, D·T and S·S (focus: Gynaecology imaging) provided valuable input into protocol review and addressed gynaecology-related issues within the manuscript. J·F·B, K·L-R, T·B, J·P, E·O·A, S·S, N·P, C·K, N·C, N·B, C·L, M·A-M, J·H·K, S·L, L·S, M·M, M·F, D·T and J·Y were all involved in revision of the manuscript and approved the final version for submission. T·B and E·O·A are the guarantors for this paper and accept full responsibility for the work and/or the conduct of the study.

## Acknowledgements

The authors acknowledge support from Medical Research Council Grant (MR/M015858/1), Imperial College NIHR Biomedical Research Centre award (WSCC_P62585), Imperial College Experimental Cancer Medicines award (C1312/A25149) and National Cancer Imaging Translational Accelerator (C2536/A28680) and Imperial Health Charity. We would also like to acknowledge the study participants, who engaged with our vision to improve the ultrasound classification of adnexal masses, through the integration of ML.

